# An analysis of buprenorphine distribution patterns among pharmacies and hospitals in the United States from 2019-2023

**DOI:** 10.1101/2024.10.02.24314657

**Authors:** Maria Gikoska, Anna K. Florio, Andrew George, Brian J. Piper

## Abstract

**Background:** Opioid Use Disorder (OUD) is a debilitating condition characterized by the overuse of prescription opioid medications and the development of physical and/or psychological dependence. Consequences of this condition include chronic impairment, distress, and later life-altering health conditions such as overdose, all of which have been highlighted by the prominence of OUD in the United States in recent years. Buprenorphine is a standard OUD treatment and commonly used for pain management. Understanding changes in distribution patterns across the US is vital for continuing to improve outcomes for OUD patients.

**Methods:** This study analyzed changes in buprenorphine distribution among pharmacies and hospitals from 2019-2023 to determine temporal patterns and identify state level disparities. The Drug Enforcement Administrations’ Automated Reports and Consolidated Ordering System (ARCOS), US Census Bureau Population Estimates databases, and mortality rates (CDC WONDER) were analyzed. Data were corrected for population to identify patterns of buprenorphine distribution in the US from 2021-2022 and 2022-2023 through examining percent changes in milligrams per 100 population at national and state level.

**Results:** The year-to-year percent change of national buprenorphine distribution from pharmacies has remained positive but changed from 12.2% increase from 2019-2020 to a four percent increase every year from 2020-2023. From 2021-2022, there was a +4.9% increase in total grams of buprenorphine distributed to pharmacies and a 95% CI [-5.1, 14.9], with District of Columbia, South Dakota, and Nebraska outside of the 95% CI. Distribution to hospitals increased by 10.2% [-32.3, 52.7] during 2021-2022, with Hawaii, New Hampshire and Delaware being outside of 95% CI. From 2022-2023, there was an increase of +5.7% and 95% CI [-3.5, 14.9] in pharmacy distribution, with states including Washington, Rhode Island and Kansas remaining outside of the 95% CI. Hospital distribution has decreased from twenty percent between 2019-2020 to eighteen percent between 2022-2023. Changes in mortality data from 2022-2023 showed no associations with trends in buprenorphine distribution for those years.

**Conclusion:** Following increases in buprenorphine distribution during the COVID pandemic, a consistent increase has continued year-over-year in most states and the country overall by both pharmacies and hospitals. Some states (e.g. Rhode Island, Georgia, Washington D.C.) have not followed this pattern. Notably, Hawaii went from the most negative percent change in hospital distribution to the most positive change in the timeframe analyzed. This may offer opportunities to analyze more specific impacts of the increased buprenorphine distribution on populations and their outcomes associated with OUD.

## 1 Introduction

Opioid Use Disorder (OUD), defined in the DSM 5 as a “problematic pattern of opioid use leading to clinically significant impairment or distress”, is a chronic and life-altering health condition that can lead to comorbidities and mortality, often due to overdose.^1^From 2018 to 2023, opioid-related overdose deaths had continued to increase in the United States (US), although in 2023, deaths decreased to an estimated 81,083 from an estimated 84,181 in 2022.^2^ While this trend appears encouraging for existing treatments and programs, approximately 5.7 million people (2.0%) were estimated to have OUD in 2023.^3^ The overall economic cost of OUD was estimated at $1.5 trillion in 2020 and has continued to increase.^4^ The continued prevalence of OUD, the costs associated with it, and most importantly, the impact on and loss of lives make insight into treatments for OUD vital.

Medications such as buprenorphine and methadone are currently the standard for OUD treatment and pain management.^5^ Buprenorphine affects all four major opioid receptors, functioning as a mu receptor partial agonist, a kappa receptor agonist, and an antagonist to both delta receptors and nociceptin receptors.^6^ Its high affinity and low intrinsic activity on mu receptors cause a weaker opioid effect than alternative options such as methadone. Compounded with a long half-life, high potency, and lower relative potential for adverse effects, buprenorphine has been identified as an optimal treatment alternative for those presenting with withdrawal.^7^ Buprenorphine has been demonstrated to reduce opioid related deaths by up to 50% through increased retention leading to risk reduction, and when prescribed at fixed, high dosages (greater than 7 mg daily), was an effective therapy for decreasing illicit opioid use.^8^ Deaths involving buprenorphine occur less frequently than methadone.^9^ Three-quarters of doctors working in emergency departments preferred buprenorphine over methadone as an OUD treatment.^10^ Considering its status as a likely safer alternative treatment option and more widely available than methadone in the US, buprenorphine is garnering continued attention in treating OUD. ^5,11^

Buprenorphine’s use as a treatment has increased across the US in a non-homogeneous manner since its approval in 2002.^11^ Policy changes at both federal and state levels have contributed to pronounced differences across states in both availability and usage.^12^ A national-level retrospective cohort study from 2012-2015 demonstrated buprenorphine treatment was highly skewed towards White patients and those with private payer insurances or self-pay.^13^ Following the emergence of SARS-CoV2, responses to the ensuing COVID-19 pandemic by governments at the federal and state levels prioritized increasing and maintaining OUD treatment access, which was ultimately beneficial in increasing buprenorphine availability and usage in the early stages.^12^

Healthcare impacts from both the pandemic and the years following are still unfolding and yet to be completely understood. This study’s objective was to compile and analyze data for buprenorphine usage by state across the US to quantify and characterize these disparities in access and distribution of buprenorphine for 2019-2023 overall as well as more focused analysis to compare patterns between 2021-2022 and 2022-2023.

## 2 Methods

### 2.1 Procedures

The annual distributions of buprenorphine were extracted from Automated Reports and Consolidated Ordering System (ARCOS) Retail Drug Summary Reports generated by the Drug Enforcement Administration (DEA).^14^ This reporting system monitors commercial distribution of controlled substances from manufacturers and distributors at the retail level. Annual estimates of the resident population were obtained from the US Census Bureau for 2019-2023.^15^ The quantities of buprenorphine (in grams) including fifty states and the District of Columbia, were obtained from the ARCOS Report 5 statistical summary for retail drug purchases for 2019 to 2023. For this analysis, our focus was to identify patterns of buprenorphine distribution (total grams) to pharmacies and hospitals post-COVID-19 pandemic and compare 2021-2022 and 2022-2023. In addition, distribution from 2019-2023 was utilized to identify an overall change in buprenorphine.

Data on mortality rates were extracted from the CDC WONDER database of public health records.^16^ Relevant ICD-10 codes for opioid overdose deaths were referenced from a prior study conducted by the Kaiser Family Foundation (KFF), a full list of which can be found in Table 1.^17^ Provisional mortality rates were queried from the CDC WONDER database by indicating multiple causes of death, including any ICD-10 designations from Group 1 (manner of death) and any ICD-10 designations from Group 2 (agent of death). Age-adjusted mortality rates by state and year were ultimately extracted, focusing on provisional data from 2022 and 2023.

### 2.2 Data Analysis

The distribution rates of buprenorphine in milligrams per 100 population were calculated for each state, overall, from 2019-2023 and annually between 2021-2022 and 2022-2023. Percentage changes in buprenorphine distribution were calculated using the formulas:

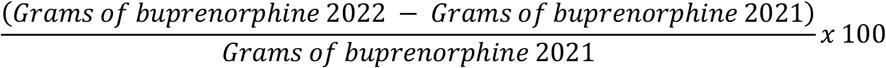

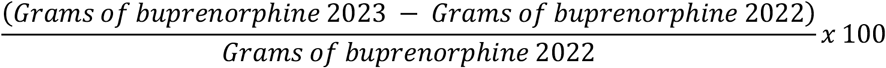

for 2021-2022 and 2022-2023, respectively. The absolute changes in buprenorphine distribution were determined by calculating the differences in grams of buprenorphine per 100K population^14^ reported from 2021-2022 and 2022-2023 and reported in units of mg/1oo people. For the calculation of 95% confidence intervals (CIs), standard deviation (SD) was determined by the STDEV.P function in Excel. The margin of error (MoE) was calculated using a= 0.05 by the equation:

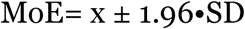

Data were plotted using GraphPad Prism and statewide geographic maps using external Heat Map application, Datawrapper.^18^

Similarly, percentage changes in age-adjusted mortality rates of opioid overdose were calculated using the formula:

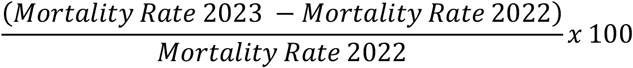

Data were plotted using GraphPad Prism.^19^

Z-scores were standardized in relation to each data set’s calculated mean (µ) and standard deviation (σ) with the following formula:

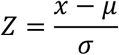

P-values were likewise calculated in Excel, with significance determined via a two-tailed z-test when α < 0.05.

Correlation determined in GraphPad Prism by calculating the r2 value using sum of squares with the following formula, where SSreg is the sum-of-squares of the regression and SStot is the sum-of-squares of the null hypothesis:

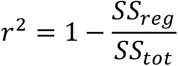

## 3 Results

### 3.1 ARCOS

At the national level, the year-to-year percent increase in buprenorphine distribution from pharmacies remained positive, though it shifted from a 12.2% rise between 2019 and 2020 to more moderate increases annually. From 2020 to 2021, distribution grew by 4.7%, followed by a 4.9% increase from 2021-2022. In addition, a 5.7% increase was observed from 2022-2023. However, the percent change of buprenorphine distribution from hospitals was +20.3% from 2019 to 2020 and +18.7% from 2022 to 2023. From 2021-2022, there was a +4.9% increase in total grams of buprenorphine distributed to pharmacies and a 95% CI [-5.1, 14.9], with District of Columbia, South Dakota and Nebraska outside of the 95% CI. Distribution to hospitals increased by 10.2% [-32.3, 52.7] during 2021-2022, with Hawaii, New Hampshire and Delaware being outside of 95% CI. From 2022-2023, there was an increase of +5.7% and 95% CI [-3.5, 14.9] in pharmacy distribution, with states including Washington, Rhode Island and Kansas remaining outside of the 95% CI. A +18.7% increase, with a 95% CI [-40.8, 78.3] was observed in distribution to hospitals, with New Hampshire and Hawaii outside of the 95% CI. The states with the largest changes from 2021-2022 in pharmacy distributions were Washington D.C (−16.62%) and Nebraska (17.1%), with only five total states decreasing. The largest changes in hospital distributions were observed in Hawaii (−55.6%) and Delaware (85.7%), with ten states displaying a decrease. The largest changes from 2022-2023 in pharmacy distributions were in Washington D.C (−7.5%) and Kansas (18.3%), with five states decreasing year-to-year. The largest changes in hospital distributions were in Maine (−16.8%) and Hawaii (188.3%) with eight states displaying a decrease. Furthermore, Washington DC maintained the smallest percent change among pharmacy distribution (−16.6% and -4.7%) from 2021-2022 and 2022-2023.

**Figure 1:**
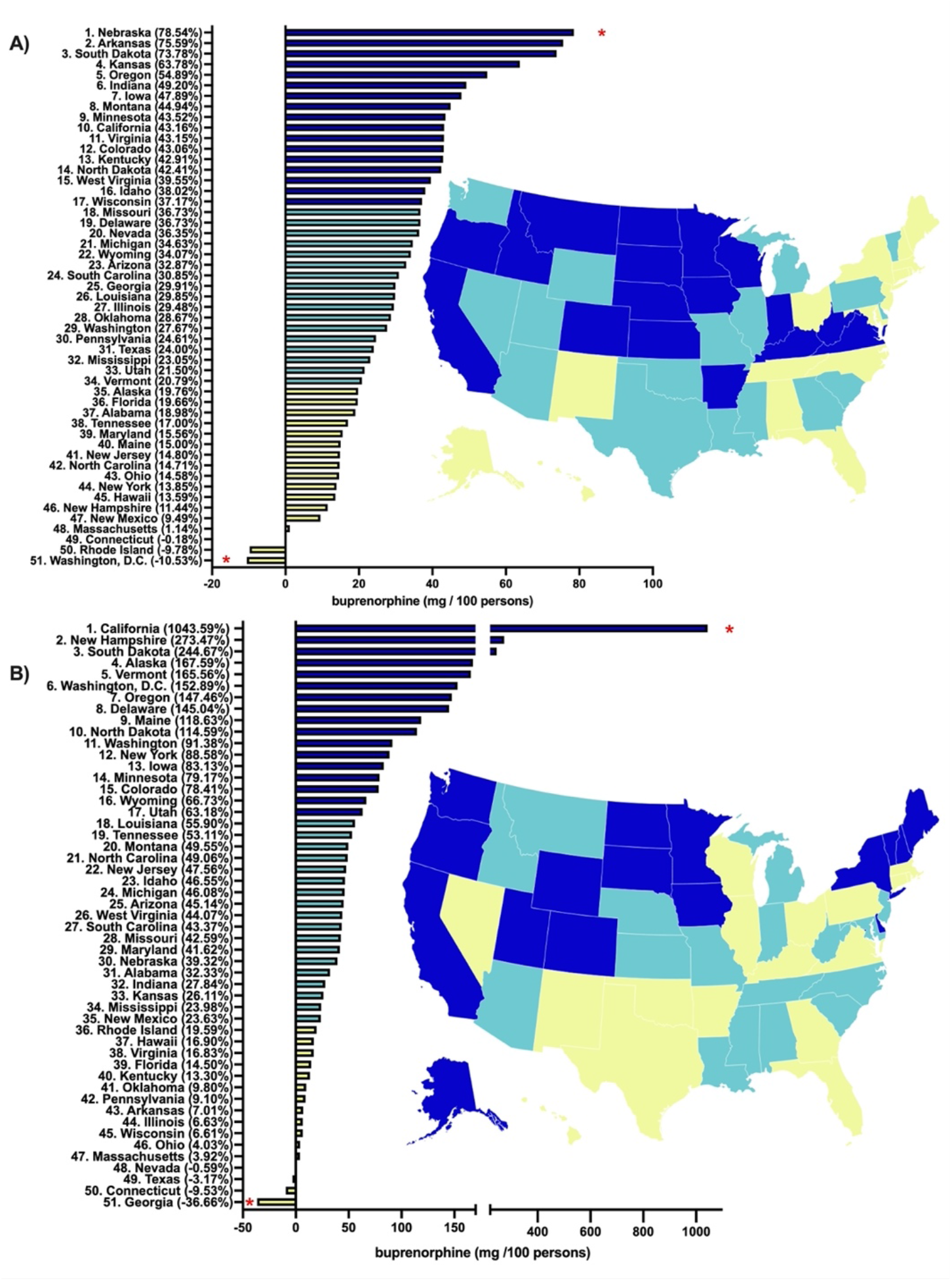
Percent change from 2019-2023 in pharmacies **(A)** and hospitals **(B)** in buprenorphine distribution as reported to the Drug Enforcement Administration’s Automated Reports and Consolidated Orders System. States outside a 95% confidence interval are designated with *.

**Figure 2:**
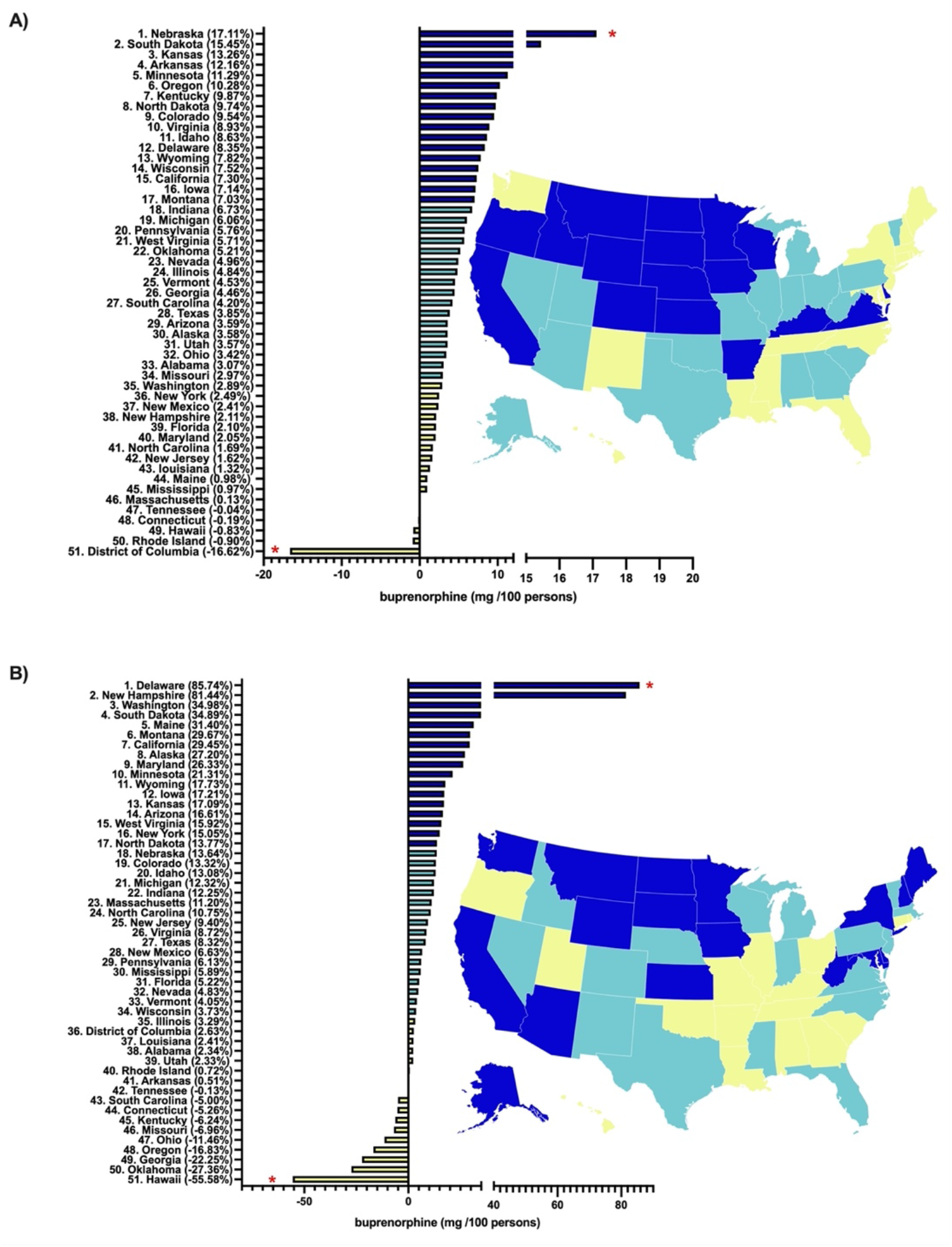
Percent change from 2021 to 2022 in pharmacies **(A)** and hospitals **(B)** in buprenorphine distribution as reported to the US Drug Enforcement Administration’s Automated Reports and Consolidated Orders System.

**Figure 3:**
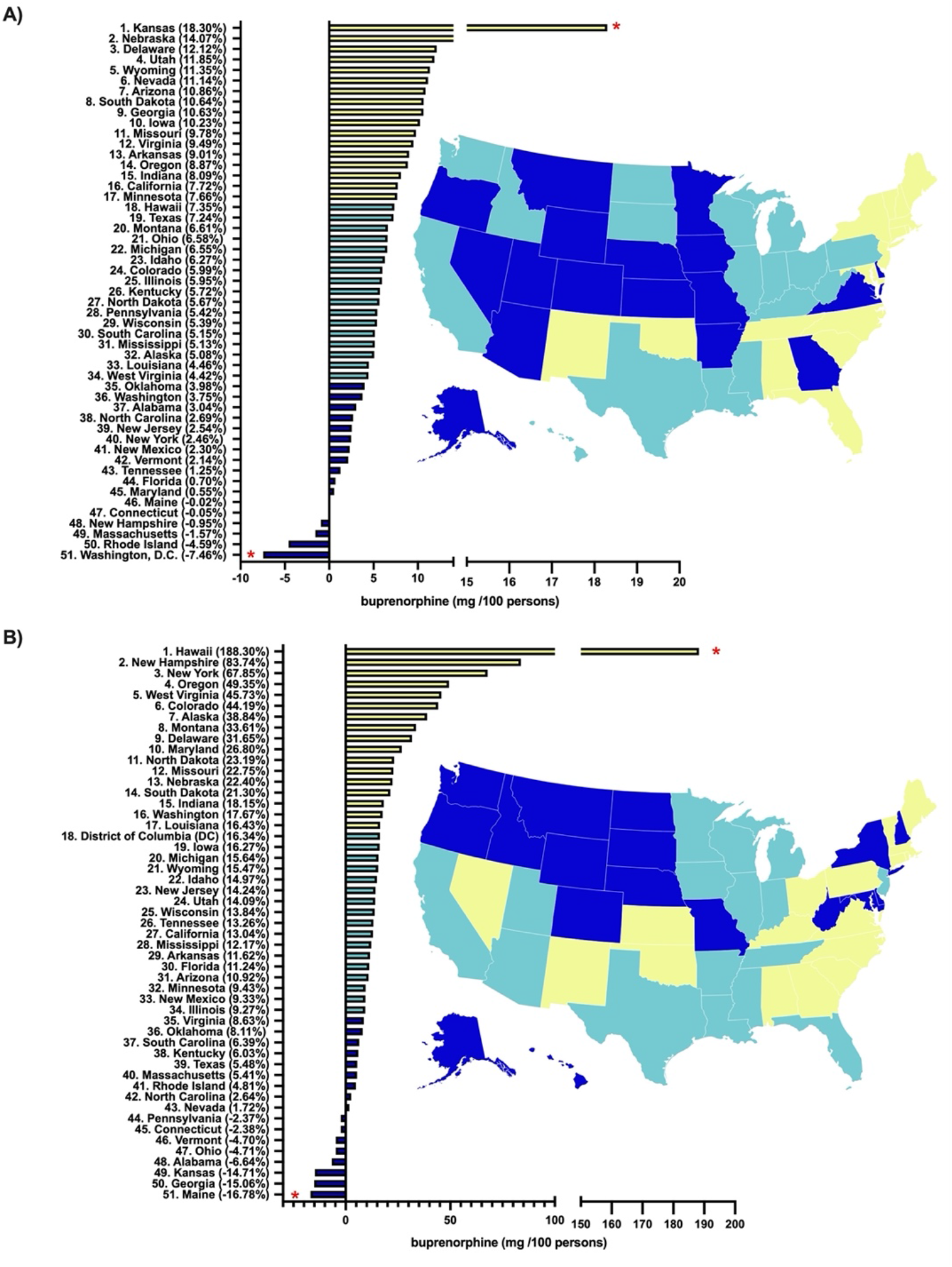
Percent change 2022-2023 in pharmacies **(A)** and hospitals **(B)** in buprenorphine distribution as reported to the US Drug Enforcement Administration’s Automated Reports and Consolidated Orders System.

### 3.2 Opioid Mortality Rates

Across all states and the District of Columbia, mortality attributed to opioids decreased from 2022-2023, with a -29.4% decrease on average and a 95% CI [-8.6, 50.2]. States with a more pronounced mortality rate reduction included Nebraska (−50.7%), North Carolina (−52.0%), and Vermont (−51.6%). States with a less pronounced mortality rate reduction included Alaska (−3.5%), Oregon (−0.8%), and Washington (−5.0%). All six of these states lie outside the confidence interval with a p-value less than 0.5. When plotted against one another, no significant correlation was observed between the percent changes of buprenorphine distribution rates in pharmacies and mortality rates from opioids in 2022-2023 (R2 value = 0.00009575, Figure 4).

**Figure 4:**
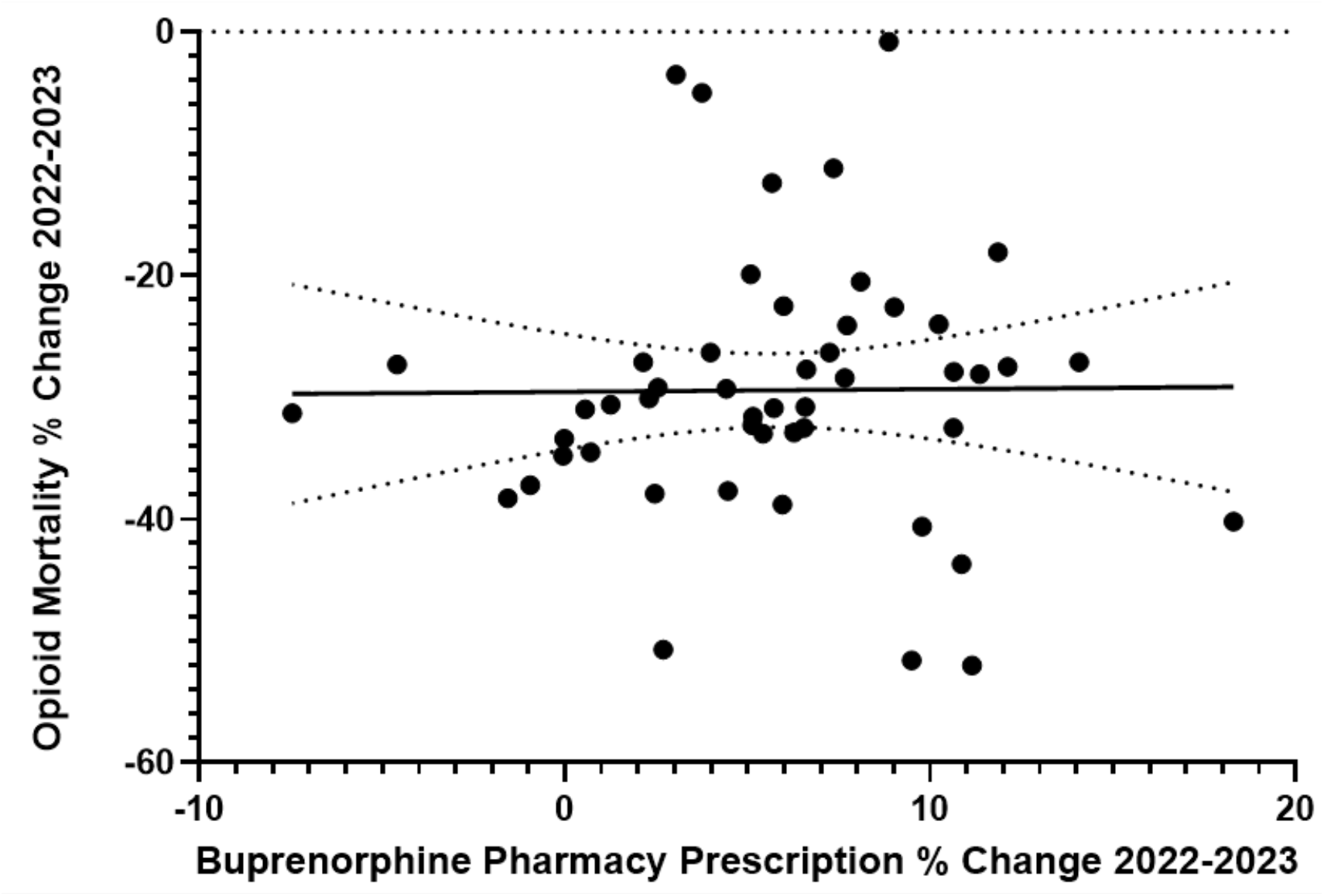
Opioid mortality percent change 2022-2023 versus the pharmacy distribution percent change (with trendline and 95% confidence interval). Equation: y = 0.02189x - 29.55 | r2 = 0.00009575

## 4 Discussion

This report identified a notable increase in buprenorphine distribution during the COVID-19 pandemic, which has consistently risen year-to-year in most states and the country overall by both pharmacies and hospitals. Analysis of ARCOS usage data revealed that in comparison to increased changes observed during COVID-19 pandemic, the percent change of buprenorphine distribution from hospitals decreased in growth to under ten percent from 2022-2023.^14^ Additionally, although it remains positive, distribution from pharmacies changed from over a ten percent increase from 2019-2020 to an increase by four percent every year from 2020-2023.^14^

Buprenorphine distribution rates to pharmacies have shown an upward pattern since the onset of COVID-19 pandemic. Initially, the increase was substantial, with growth rates exceeding ten percent from 2019 to 2020. The initial rise in distribution can be attributed to increased demand for substance use treatment related to stressors and disruptions in access to traditional treatment settings related to the pandemic.^13^ Additionally, changes in regulations and services, such as telehealth, contributed to the sharp increase, promoting easier access to medications like buprenorphine.^20,21^ However, as the pandemic progressed, the growth rate stabilized around four percent per year suggesting an ongoing but moderate expansion in buprenorphine availability to pharmacies reflecting emergency related increases to more stable growth patterns. This moderation in buprenorphine distribution rate may be due to factors including reaching a plateau in buprenorphine demand, adaptation of treatment systems, and supply chain constraints.^22,23^

In contrast, the distribution rates of buprenorphine to hospitals were observed to increase during the COVID-19 pandemic as hospitals acclimated to the need for emergency care and the expansion of treatment services exacerbated by the pandemic.^21^ The decrease in buprenorphine distribution from hospitals observed from 2022 to 2023 can be attributed to several factors such as decrease in buprenorphine demand as pandemic subsided and reduced supply of medications within hospital settings.^24^ Additionally, hospitals focus on acute and stabilizing care in comparison to long-term maintenance managed through outpatient settings, explaining the more consistent increases observed in pharmacy distribution due to the integration of routine outpatient care.^13, 25^

Some states, such as Rhode Island, Georgia, and Washington D.C., have not followed this pattern. Strikingly, in the timeframe analyzed, Hawaii began with the most negative percent change in hospital buprenorphine distribution but finished with the most positive. These abnormalities interestingly coincided with a few notable legislative and administrative actions during the COVID-19 pandemic to address OUD. In Washington D.C, expansion of telemedicine services allowed for remote prescribing and increased distribution of buprenorphine during the COVID-19 lockdown.^26^ Similar policy changes reduced barriers to accessing treatment (i.e. clinic closure and infective precautionary measures), and could explain increased buprenorphine prescriptions during the pandemic.^20,21^ In contrast, Hawaii experienced more restricted access to buprenorphine due to limited expansion of telemedicine services and distribution of the quantity of medication prescribed without sufficient visits, resulting in more gradual changes over time.^23,24^ This could have led to reduced buprenorphine treatment during the pandemic.^27^

The increase in buprenorphine distribution is a positive indicator of increased demand for the medication. Despite the rise in buprenorphine availability, overdose rates remain a concern.^28^ States have shown a general decrease in mortality rates from 2022-2023, which could indicate widespread progress with regards to reducing the impacts of OUD in the United States. Considering no trend could be established between reduced mortality and changes in buprenorphine pharmacy prescription, however, there is no evidence to suggest that increased access to buprenorphine alone would be the sole cause of that reduction. Larger scale analysis of factors contributing to reduced mortality rates from opioids may be required to better understand which improvements have thus far been effective, which changes have failed to be effective, and which areas can be focused on in the future to further reduce mortality, among other complications.

The effectiveness of buprenorphine for OUD treatment and reducing opioid overdose deaths has been well investigated, highlighting the need for strategies that include medication assisted treatment and public health interventions.^29^ Future studies may be indicated to further analyze associations between buprenorphine distributions and state-level policy changes, specifically regarding proactive versus restrictive approaches, with the aim to expand access to OUD treatments. Additionally, research should explore the integration of buprenorphine with other interventions such as mental health services, facilitating a more holistic approach to combating opioid overdoses. Identifying and addressing factors impacting buprenorphine availability and effectiveness in diverse populations will be crucial for improving its role in response to the opioid crisis.

The method of buprenorphine distribution reporting in ARCOS introduces potential limitations to this study. First, ARCOS reports buprenorphine weight rather than prescriptions per person, which can lead to inaccuracies if weights vary significantly for each prescription. The reporting system also does not differentiate between mono- and combo products, so it remains unclear which formulations are more available, encouraged, or effective. Furthermore, ARCOS lacks detailed patient information revealing medical comorbidities or social determinants of health, limiting our understanding of the treatment scope or any confounding factors. Future studies may be conducted to examine relations between social determinants of health and availability of treatments for OUD (like buprenorphine).

Note that this study analyzed state-level patterns, and we recognize the vast diversity and variation within each state cannot be fully appreciated within this study’s limited scope. Future studies regarding economic or cultural variations within a smaller region may be warranted to better understand socioeconomic contributors to opioid treatment. Further expansion of research may also consider comparing drug rehabilitation admissions and recovery.

## Data Availability

All data produced in the present study are available upon reasonable request to the authors
All data produced in the present work are contained in the manuscript

## ACKNOWLEDGMENTS

This research received no external funding. MG, AF, AG conducted data gathering and analysis. MG conducted literature review, statistical analyses of annual distribution rates of buprenorphine to pharmacies and hospitals per population and created figures. AF conducted analysis of mortality data, literature review and created figures. AG conducted literature review and editing of statistical analyses. MG, AF, AG and BP contributed to writing and editing of the manuscript. BJP was (2019-21) part of an osteoarthritis research team supported by Pfizer and Eli Lilly.

## Appendix

**Table 1:** ICD-10 Codes Referenced for CDC WONDER Provisional Mortality Data.

|  | ICD-10 Codes | Causes of Death |
| --- | --- | --- |
| Group 1 | X40 | Accidental poisoning by and exposure to nonopioid analgesics, antipyretics and antirheumatics |
|  | X41 | Accidental poisoning by and exposure to antiepileptic, sedative-hypnotic, antiparkinsonism and psychotropic drugs, not elsewhere classified |
|  | X42 | Accidental poisoning by and exposure to narcotics and psychodysleptics [hallucinogens], not elsewhere classified |
|  | X43 | Accidental poisoning by and exposure to other drugs acting on the autonomic nervous system |
|  | X44 | Accidental poisoning by and exposure to other and unspecified drugs, medicaments and biological substances |
|  | X60 | Intentional self-poisoning by and exposure to nonopioid analgesics, antipyretics and antirheumatics |
|  | X61 | Intentional self-poisoning by and exposure to antiepileptic, sedative-hypnotic, antiparkinsonism and psychotropic drugs, not elsewhere classified |
|  | X62 | Intentional self-poisoning by and exposure to narcotics and psychodysleptics [hallucinogens], not elsewhere classified |
|  | X63 | Intentional self-poisoning by and exposure to other drugs acting on the autonomic nervous system |
|  | X64 | Intentional self-poisoning by and exposure to other and unspecified drugs, medicaments and biological substances |
|  | X85 | Assault by drugs, medicaments and biological substances |
|  | Y10 | Poisoning by and exposure to nonopioid analgesics, antipyretics and antirheumatics, undetermined intent |
|  | Y11 | Poisoning by and exposure to antiepileptic, sedative-hypnotic, antiparkinsonism and psychotropic drugs, not elsewhere classified, undetermined intent |
|  | Y12 | Poisoning by and exposure to narcotics and psychodysleptics [hallucinogens], not elsewhere classified, undetermined intent |
|  | Y13 | Poisoning by and exposure to other drugs acting on the autonomic nervous system, undetermined intent |
|  | Y14 | Poisoning by and exposure to other and unspecified drugs, medicaments and biological substances, undetermined intent |
| Group 2 | T40.0 | Opium |
|  | T40.1 | Heroin |
|  | T40.2 | Other opioids |
|  | T40.3 | Methadone |
|  | T40.4 | Other synthetic narcotics |
|  | T40.6 | Other and unspecified narcotics |

## Supplemental Figures

**Figure 4:**
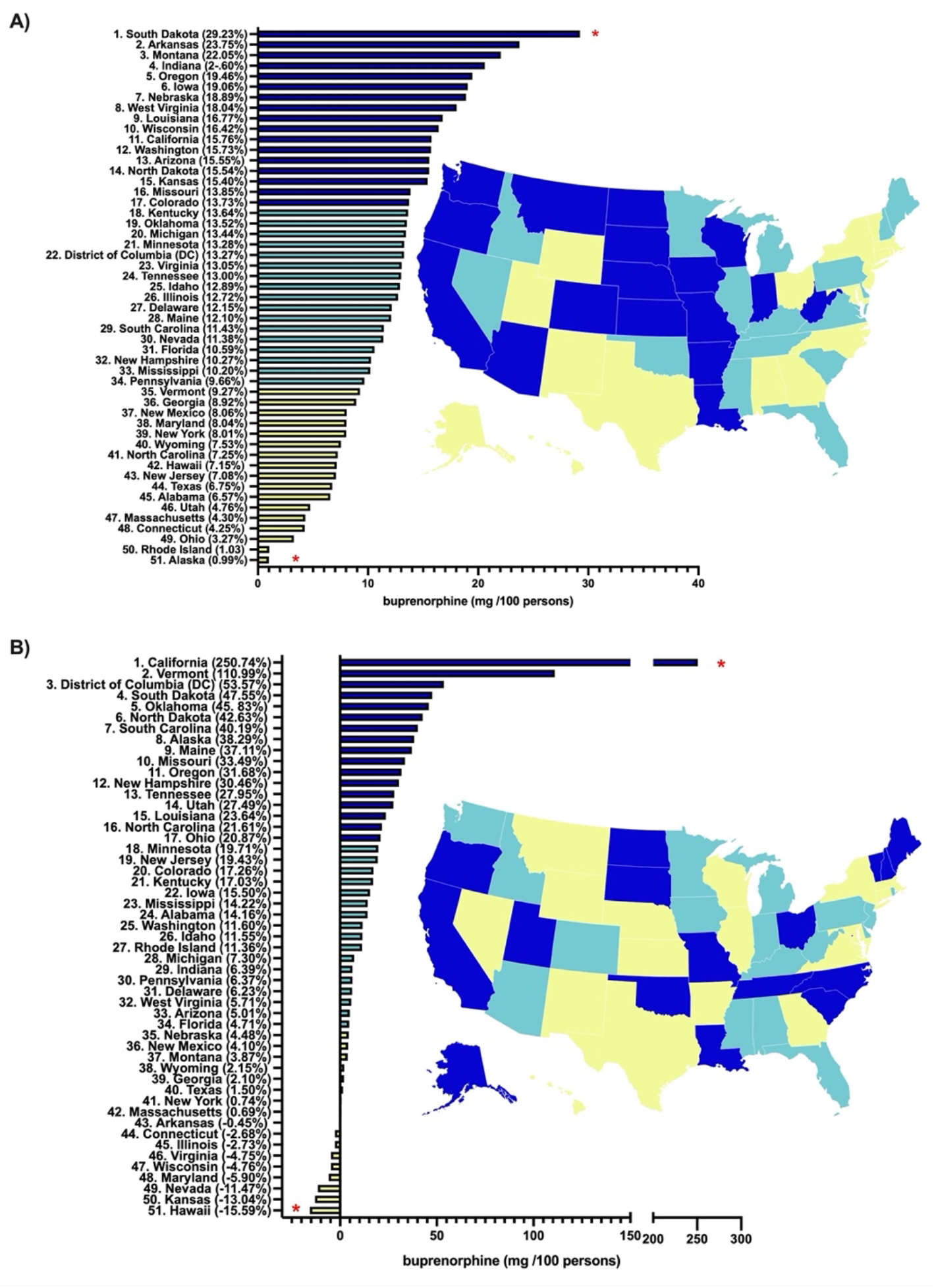
Percent Change 2019-2020 Pharmacies **(A)** and hospitals **(B)** in buprenorphine distribution as reported to the Drug Enforcement Administration’s Automated Reports and Consolidated Orders System.

**Figure 5:**
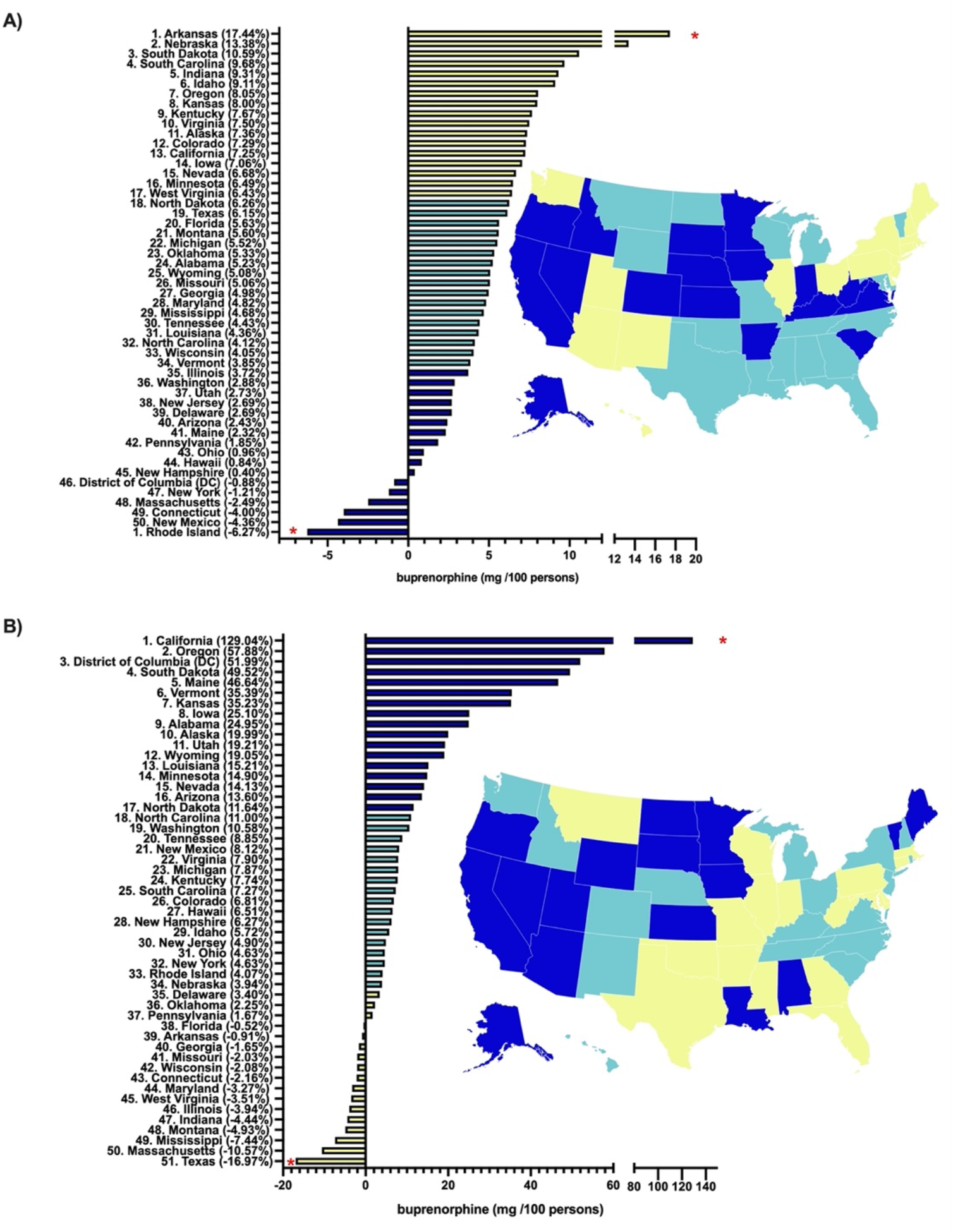
Percent Change 2020-2021 Pharmacies **(A)** and hospitals **(B)** in buprenorphine distribution as reported to the Drug Enforcement Administration’s Automated Reports and Consolidated Orders System.

## Notes

### Competing Interest Statement

The authors have declared no competing interest.

### Funding Statement

This study did not receive any funding

### Summary of Updates

Abstract section: Background; included pain management.

